# Severe COVID-19 induces molecular signatures of aging in the human brain

**DOI:** 10.1101/2021.11.24.21266779

**Authors:** Maria Mavrikaki, Jonathan D. Lee, Isaac H. Solomon, Frank J. Slack

## Abstract

Coronavirus disease 2019 (COVID-19) is predominantly an acute respiratory disease caused by severe acute respiratory syndrome coronavirus 2 (SARS-CoV-2) and remains a significant threat to public health. COVID-19 is accompanied by neurological symptoms and cognitive decline, but the molecular mechanisms underlying this effect remain unclear. As aging induces distinct molecular signatures in the brain associated with cognitive decline in healthy populations, we hypothesized that COVID-19 may induce molecular signatures of aging. Here, we performed whole transcriptomic analysis of human frontal cortex, a critical area for cognitive function, in 12 COVID-19 cases and age- and sex-matched uninfected controls. COVID-19 induces profound changes in gene expression, despite the absence of detectable virus in brain tissue. Pathway analysis shows downregulation of genes involved in synaptic function and cognition and upregulation of genes involved in immune processes. Comparison with five independent transcriptomic datasets of aging human frontal cortex reveals striking similarities between aged individuals and severe COVID-19 patients. Critically, individuals below 65 years of age exhibit profound transcriptomic changes not observed among older individuals in our patient cohort. Our data indicate that severe COVID-19 induces molecular signatures of aging in the human brain and emphasize the value of neurological follow-up in recovered individuals.

## Introduction

Although COVID-19 is primarily a respiratory disease, neurological symptoms are also reported in a subpopulation of infected COVID-19 individuals [1], with substantially higher rates (up to 84%) in severe COVID-19 cases [2]. For instance, patients with prior severe COVID-19 exhibit a 10-year average drop in their global cognitive performance [3]. Complementary studies combining neuroimaging and cognitive screening implicate COVID-19-induced impairment of the frontal cortex [4, 5], a critical area for cognitive function [6, 7]. Furthermore, the effects of COVID-19 on the nervous system, including cognitive impairment, are expected to be long-term [3, 8-10]. However, the molecular mechanisms underpinning the effects of COVID-19 on cognition remain to be determined.

In healthy populations, the natural process of aging leads to a reduction in frontal cortex activity [11] and cognitive decline [12]. At the molecular level, aging induces distinct molecular signatures in the human brain, including increased activation of immune signaling and decreased synaptic activity [13-15]. Based on the deteriorating effects of COVID-19 on the frontal cortex and on cognitive function, paralleling the effects of aging, we hypothesized that COVID-19 induces molecular signatures of aging in the frontal cortex.

### COVID-19 gene expression in human brain

To test this hypothesis, we performed whole transcriptomic analyses of postmortem frontal cortex of 12 severe COVID-19 patients (with pre- or peri-mortem positive testing for SARS-CoV-2 by nasopharyngeal swab qPCR and history of hospitalization) and 12 age- (±2 years) and sex-matched uninfected controls (**Fig. 1a**). Clustering analysis via t-distributed stochastic neighbor embedding (TSNE) reveals separation of COVID-19-associated transcriptomic profiles from control profiles (**Fig. 1b**), with the two controls proximal to COVID-19 cases being from elderly patients (ages 71 and 84). To assess whether this clustering is due to the presence of SARS-CoV-2 virus, we tested frontal cortex samples by quantitative PCR (qPCR) using two different primer sets recommended by the Center for Disease Control and Prevention (**Fig. 1c, Supplementary Fig. 1a**). No signal was detected above background noise for either of these primer sets. Bioinformatic alignment of sequencing reads to the SARS-CoV-2 genome also did not detect viral reads in COVID-19 or control samples (**Supplementary Fig. 1b**). In agreement with previous studies [16, 17], our data show absence of detectable virus in the frontal cortex at the time of death and suggest that the observed gene expression changes are not due to direct effects of SARS-CoV-2 virus in the frontal cortex.

**Fig 1.**
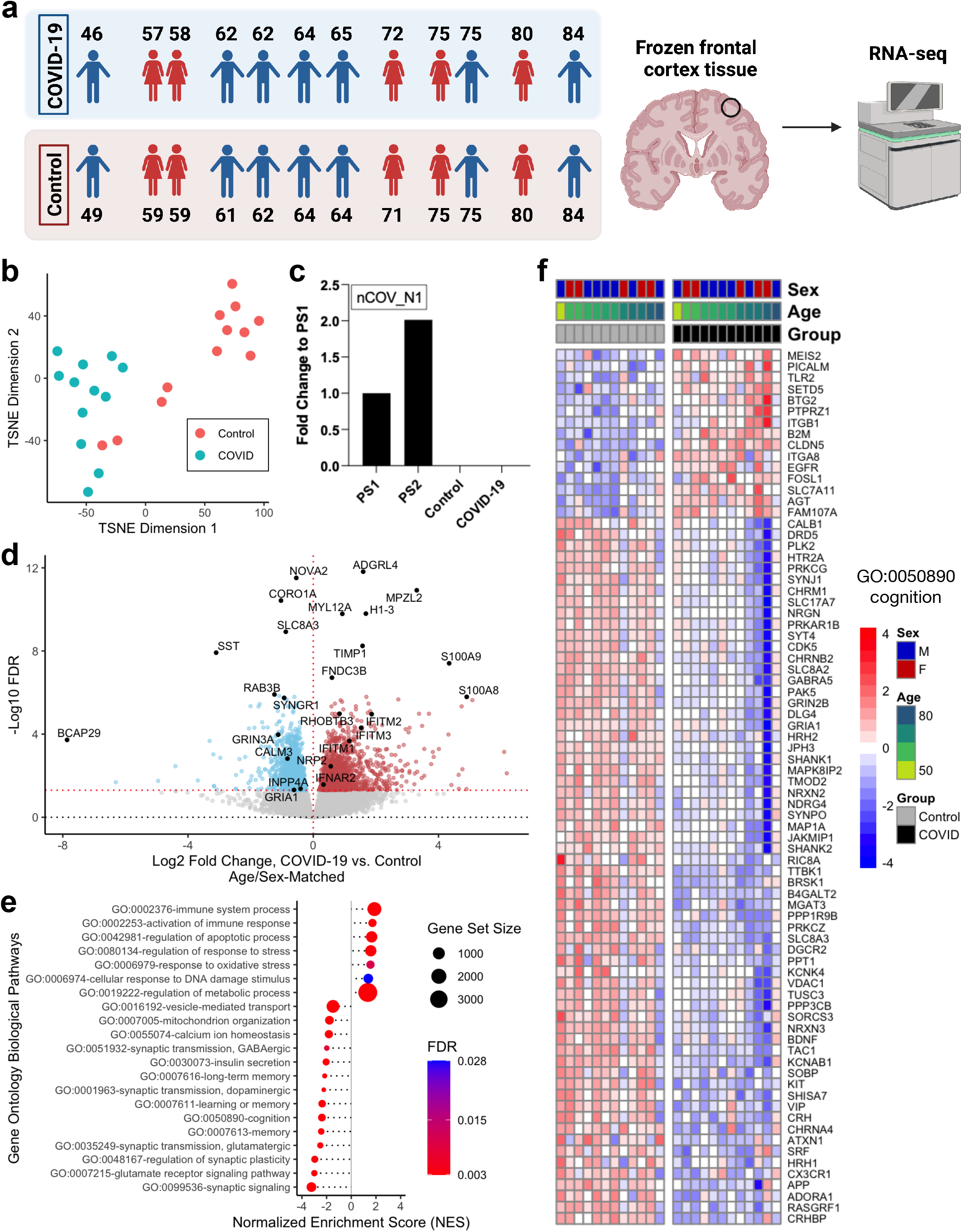
Severe COVID-19 induces global transcriptomic changes in the frontal cortex of the brain. **a**. Left, age and sex of each individual in COVID-19 or control groups (n=12/group) analyzed in this cohort. Each COVID-19 case was matched with an uninfected control case by sex and age (±2 years). Right, schematic of study approach. *Schematic was generated with BioRender*. **b**. t-distributed stochastic neighbor embedding (TSNE) analysis of frontal cortex transcriptomes from COVID-19 cases and uninfected controls. **c**. qPCR assessment of SARS-CoV-2 viral RNA in the frontal cortex using the nCOV_N1 primer set. PS1 and PS2 correspond to the 2019-nCoV_N_Positive Control RUO Plasmid (IDT) at concentrations of 1,000 and 2,000 copies/μl, respectively (a technical duplicate/concentration was used to estimate the corresponding mean; for control and COVID-19 samples n=13/group). **d**. Volcano plot representing the differentially expressed genes (DEGs) of the frontal cortex of COVID-19 cases versus matched controls. Red points, significantly upregulated genes among COVID-19 cases (false discovery rate < 0.05). Blue points, significantly downregulated genes among COVID-19 cases. Black points, highlighted significant genes with corresponding gene symbols. **e**. Gene ontology (GO) biological pathway enrichment analysis of COVID-19 versus control brain DEGs. Gene ranks were determined by signed -log10 false discovery rates of DEGs. FDR, gene set enrichment analysis false discovery rate. **f**. Heatmap of relative gene expression levels of significant DEGs associated with the “cognition” (GO: 0050890) GO term across COVID-19 and control samples. Color legend, scaled gene expression levels across subjects, normalized via variance-stabilized transformation.

We next compared COVID-19 cases to their corresponding age- and sex-matched controls and identified a total of 2,809 differentially expressed genes (DEGs) with unique Ensembl gene IDs, 1,397 of which were upregulated and 1,412 downregulated (**Fig. 1d, Supplementary Fig. 2a-b**). In the human brain, we did not observe expression of genes involved in SARS-CoV-2 entry, such as *ACE2* or *TMPRSS2* (**Supplementary Fig. 2c**) [8, 16]. On the other hand, *S100A8*/*S100A9* genes, which encode for calprotectin and circulating levels of which distinguish severe from mild COVID-19, were upregulated among COVID-19 patients in our cohort [18, 19]. In addition, *SYNGR1*, previously found to be downregulated in astrocytes and monocytes/macrophages in the prefrontal cortex of severe COVID-19 patients [17], was also downregulated in our cohort. Taken together, our analysis recapitulates many gene expression findings previously associated with COVID-19.

To determine the functional roles of COVID-19 transcriptome-wide changes, we performed pathway enrichment analysis using annotated Gene Ontology (GO) Biological Processes (**Fig. 1e**). To our surprise, we found numerous significant DEGs and GO terms associated with aging in the human brain to be significantly enriched upon severe COVID-19. In agreement with previous studies [16, 17], we observed positive enrichment of immune response terms (GO:0002376; GO:0002253) together with many immune-related genes upregulated in our cohort (e.g., *BCL2, IFI16*, and *CFH*) (**Fig. 1e** and **Supplementary Fig. 3**). Of the top dysregulated genes, we observed significant increases in *IFITM1-3* gene expression levels, which are associated with interferon response [20]. Furthermore, we identified numerous synaptic function terms to be negatively enriched, including synaptic signaling (GO:0099536), regulation of synaptic plasticity (GO:0048167), and glutamatergic (GO:0035249), GABAergic (GO:0051932), and dopaminergic (GO:0001963) synaptic transmission, with corresponding downregulation of synaptic signaling genes (*SST, GRIA1*, and *GRIN2B*) (**Fig. 1e** and **Supplementary Fig. 3**). Indeed, *SST*, one of the most downregulated genes in our COVID-19 cohort, has been previously associated with aging in the human frontal cortex [14, 21]. We also observed significant associations of cellular response to DNA damage (GO:0006974) [14, 22], mitochondrial function (GO:0007005), regulation of response to stress (GO:0080134) and oxidative stress (GO:0006979), vesicular transport (GO:0016192), calcium homeostasis (GO:0055074) [14], apoptosis (GO:0042981) [23], and insulin signaling (insulin secretion: GO:0030073) [24-26] pathways previously associated with aging processes and brain aging [14, 21]. Critically, numerous COVID-19 DEGs are involved in cognitive function (cognition: GO:0050890; memory: GO:0007613, GO:0007616; and learning or memory: GO:0007611). Assessment of the DEGs overlapping with these enriched pathways reveals genes such as the brain-derived neurotrophic factor (*BDNF)* which have been associated with aging [27, 28](**Fig. 1f**). KEGG and Reactome pathway enrichment analyses also reveal positive enrichment of immune activation and negative enrichment of synaptic function pathways (**Supplementary Fig. 4**). Interestingly, KEGG analysis identifies the “Coronavirus Disease - COVID-19” pathway, based largely on non-brain tissue effects of COVID-19, as one of the most significantly enriched pathways among COVID-19 patients (**Supplementary Fig. 4**). Altogether, our analyses suggest that many biological pathways that change with natural aging in the brain also change in severe COVID-19.

### Molecular signatures of brain aging

Based on the significant enrichment patterns of cognition pathways in COVID-19 brains, together with the numerous enriched biological processes associated with aging, we sought to test whether severe COVID-19 induces similarly widespread transcriptomic changes as aging in the human frontal cortex. We collated transcriptome-wide datasets from five independent patient cohorts assessing age-dependent changes in the frontal cortex (**Fig. 2a**). For all frontal cortex aging datasets, genes upregulated in aging are upregulated in severe COVID-19; likewise, genes downregulated in aging are also downregulated in severe COVID-19 (**Fig. 2b-c** and **Supplementary Fig. 5**). Furthermore, a gene set previously associated with brain aging is strongly upregulated in COVID-19 patients [29] (**Supplementary Fig. 6**). As additional support of this association between frontal cortex aging and severe COVID-19, we confirmed upregulation of *S100A9, MYL12A*, and *RHOBTB3* genes and downregulation of *CALM3, INPP4A, GRIA1*, and *GRIN3A* in COVID-19 patients by qPCR, genes that are also differentially expressed in aged frontal cortex (**Fig. 2d**). Thus, a substantial set of DEGs in aged frontal cortex are also differentially expressed in the frontal cortex of severe COVID-19 patients.

**Fig 2.**
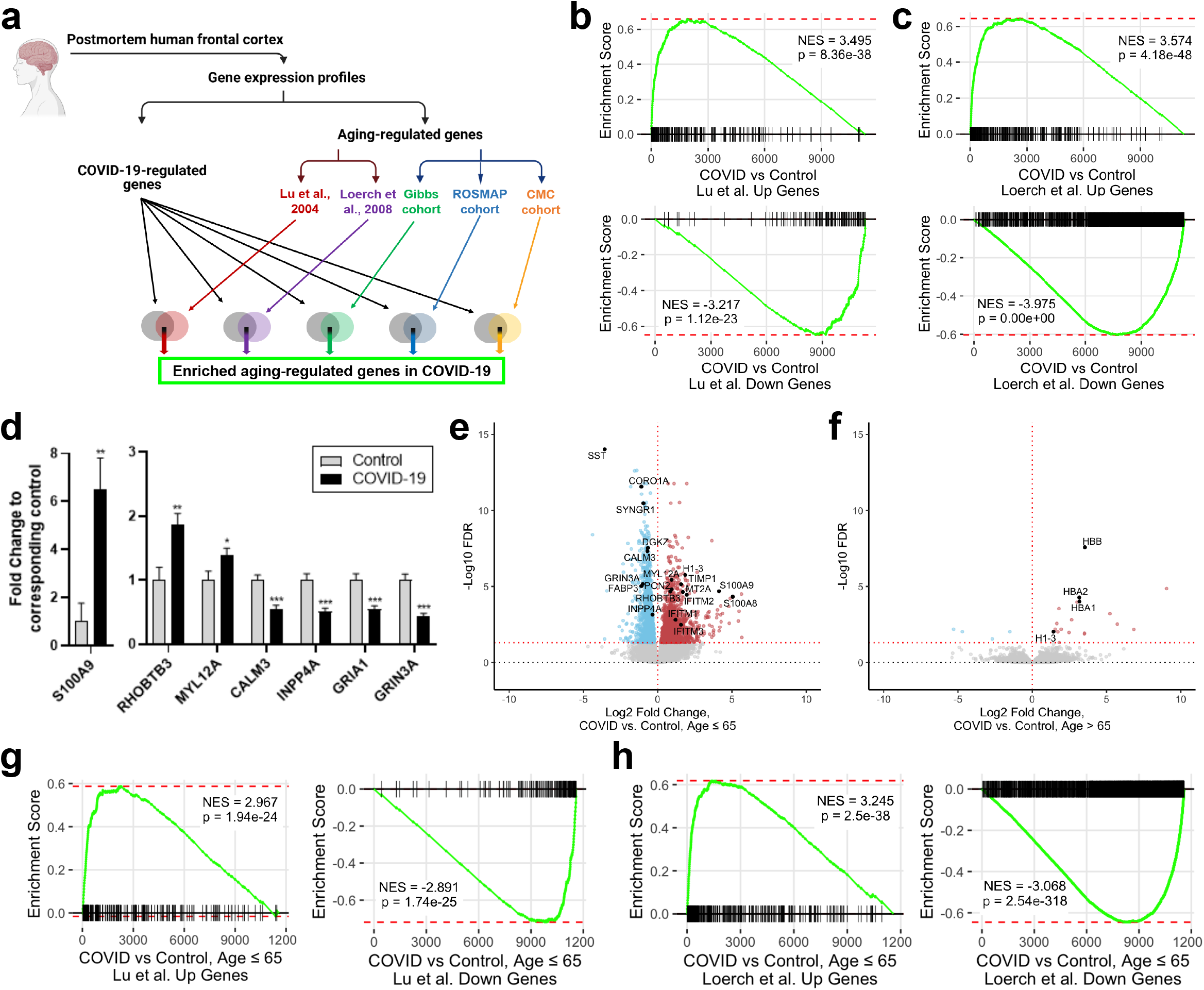
Severe COVID-19 induces transcriptomic signatures of aging in the human brain. **a**. COVID-19-associated DEGs were assessed for enrichment of brain aging DEGs curated from each of five independent patient cohort studies (Lu et al., 2004, n=21 [14]; Loerch et al., 2008, n=28 [21]; Gibbs cohort, n=37 [38, 42]; ROSMAP cohort, n=117 [40-43], Common Mind Consortium/CMC cohort, n=155 [39, 42]; n refers to the number of individuals analyzed in each cohort). *Schematic was generated with BioRender*. **b** and **c**. Gene set enrichment analysis of COVID-19 DEGs, using significantly up- (top) or down-regulated genes (bottom) in the Lu et al. (2004) (**b**) and Loerch et al. (2008) (**c**) cohorts as gene sets. DEG ranks were assigned by signed -log10 false discovery rates from COVID-19 versus control frontal cortex. NES, normalized enrichment score. p, GSEA p-value. See also **Supplementary Fig. 5. d**. qPCR assessment of candidate COVID-19 DEGs associated with aging (n=13/group). \**p*<0.05; \*\**p*<0.01; \*\*\**p*<0.001. Data are expressed as fold change relative to control ± SEM. **e** and **f**. Volcano plots representing the DEGs of the frontal cortex of COVID-19 cases versus controls among patients ≤ 65 (**e**, n=7/group) or > 65 (**f**, n=5/group) years of age. Red points, significantly upregulated genes among COVID-19 cases (false discovery rate < 0.05). Blue points, significantly downregulated genes among COVID-19 cases. Black points, significant genes with corresponding gene symbols. **g** and **h**. Gene set enrichment analysis of COVID-19 DEGs among individuals ≤ 65 years of age, using significantly up- (left) or down-regulated genes (right) in the Lu et al. (2004) (**g**) and Loerch et al. (2008) (**h**) cohorts as gene sets. DEG ranks were assigned by signed -log10 FDR values from COVID-19 versus control brains among individuals ≤ 65 years of age. NES, normalized enrichment score. p, GSEA p-value. See also **Supplementary Fig. 7**.

Next, we sought to determine if the aging gene signature in the brain might differ between younger (at or below 65 years of age, n=7/group) and older (above 65 years of age, n=5/group) patients in our study cohort. To our surprise, we observed substantial transcriptomic changes upon COVID-19 infection in the younger patient cohort, with minimal observable gene expression changes in the older cohort (**Fig. 2e-f**). In older patients, COVID-19 upregulated the expression of only 19 genes, including *HBB, HBA1* and *HBA2*, and downregulated the expression of 4 genes (**Fig. 2f**). Among younger individuals (at or below 65 years of age), we identified 1,631 upregulated genes and 2,073 downregulated genes in COVID-19, with many of the same DEGs from the age-/sex-matched full cohort also exhibiting significant differential expression. Furthermore, these DEGs retain the same significant trends with aging-associated genes in the frontal cortex (**Fig. 2g-h and Supplementary Fig. 7**). Altogether, our analyses suggest that the brain aging effects of COVID-19 are far more pronounced in younger patients than in older patients.

Finally, we tested whether the molecular changes in the brain induced by COVID-19 differ between males (n=7/group) and females (n=5/group). We observed many significant DEGs in both male (811 DEGs with unique Ensembl gene IDs; 396 were upregulated and 415 were downregulated) and female (408 DEGs; 231 were upregulated and 177 were downregulated) cohort subsets, including DEGs from our full cohort analysis (**Supplementary Fig. 8a-c**). Furthermore, both males and females exhibit concordant changes in aging-associated genes and pathways upon COVID-19 (**Supplementary Fig. 8d-e**). Thus, both males and females exhibit aging-associated transcriptomic changes in the frontal cortex due to severe COVID-19.

## Discussion

Several studies have been published assessing the transcriptomic changes induced by severe COVID-19 in the human brain [16, 17, 30]. In agreement with our findings, single cell RNA-seq studies of COVID-19 frontal cortex have identified increased immune activities [16, 17] and decreased expression of genes involved in synaptic signaling [16], together with the absence of detectable SARS-CoV-2 expression at the time of death [16, 17]. However, no study has yet demonstrated the striking and profound similarities of transcriptomic profiles between COVID-19 disease and aging in the human brain. We believe this is for two reasons: (1) our patient cohort was rigorously age- and sex-matched, and (2) the effects of COVID-19 on the brain are most substantial among patients younger than 65 years of age, multiple of who are represented in our cohort. Indeed, a recent RNA-seq study of the frontal cortex of COVID-19 individuals all above 67 years of age identified minimal transcriptomic changes (11 differentially expressed genes) [30]. Of the differentially expressed genes reported, we also observed *HBA1, HBA2*, and *HBB* genes upregulated in our older patient cohort subset (>65 years of age). Further mechanistic assessment of the brain aging-like profile observed among COVID-19 patients should thus be focused on younger patient cohorts to capture these substantial transcriptomic effects.

We recognize limitations in our study design: the variability in illness duration, the imperfect quality of several samples (as previously reported in similar studies [16]), the modest number of subjects (12 cases and 12 controls), the lack of young COVID-19 subjects, and the specificity of our findings due to COVID-19. Despite these constraints, we were sufficiently powered to identify substantial transcriptome-wide changes between COVID-19 cases and controls, including among younger patients in our patient cohort. Furthermore, in addition to being age-matched, our experimental sample size is larger than previously reported COVID-19 brain transcriptome studies [16, 17, 30], enabling the identification of aging-associated gene expression signatures in our samples. Although our study does not examine the specificity of COVID-19-induced transcriptomic changes in the brain, the implications of our findings may readily extend to related pathologies. For instance, prior clinical trials have shown that cognitive impairment is observed in 55% of survivors of severe acute respiratory syndrome (SARS) 12 months after discharge [31]. Such behavioral observations suggest that similar molecular effects in the brain may be observed not only in severe COVID-19 but also in other conditions characterized by increased peripheral and central inflammation, severe hypoxic insults, and microvascular brain pathologies [1, 32].

Aging is a major risk factor for the development of cognitive deficits and neurodegenerative diseases [12, 33, 34]. Although the molecular changes in the brain upon COVID-19 cannot be readily assessed in recovered individuals, our data herein suggest that severe COVID-19 induces premature aging in the human brain, particularly among younger individuals. Together with previously reported residual cognitive deficits observed in recovered COVID-19 individuals [3], our results imply that increased long-term rates of cognitive decline and neurodegenerative disorders may be observed among COVID-19 patients as a consequence of long COVID. In light of this possibility, we advocate for neurological follow-up of recovered COVID-19 patients and suggest potential clinical value in modifying risk factors to reduce the risk or delay the development of aging-related neurological pathologies such as dementia [35].

## Data Availability

All data produced in the present study will be available upon publication.

## Acknowledgments

This work was supported by a grant from the NIA (R01 AG058816) to F.J.S. We thank the National Institutes of Health (NIH) NeuroBioBank, the Harvard Brain Tissue Resource Center (HBTRC), and the Miami Brain Endowment Bank for providing control brain tissues. We also thank Dr. Sabina Berretta, Director of the HBTRC, for advice on the selection of appropriate brain area controls and Tanvi Saxena for advice on library preparation. We thank the BIDMC IBC and Robert Griffin for advice on BL2+ protocols, Dr. Victoria Petkova for RNA-seq library quality control, and Dr. Ioannis Vlachos for sequencing assistance.

## Author Contributions

M.M. conceived of the idea and designed the study. M.M. performed the library preparation for RNA-seq and qPCR experiments. J.D.L. performed bioinformatic analyses and generated all relevant files including codes. I.H.S. generated the relevant IRB protocol, collected COVID-19 patient brain tissues, provided clinical annotations and determined the appropriate brain area for the controls. F.J.S. and M.M. supervised the study. M.M., J.D.L. and F.J.S. wrote the manuscript. All authors reviewed and edited the manuscript prior to submission.

## Declarations of interests

The authors declare no conflicts of interest.

## Methods

### Human brain tissues

Post-mortem brain tissue specimens from individuals with severe COVID-19 were collected through an excess tissue waived consent protocol approved by the Mass General Brigham Institutional Review Board. Consent for autopsy was provided by the patients’ next of kin or healthcare proxy per Massachusetts state law. All autopsies were performed at Brigham and Women’s Hospital from 9/1/2020 to 3/30/2021 with pre- or peri-mortem positive testing for SARS-CoV-2 by nasopharyngeal swab qPCR. COVID-19 cases had no known psychiatric or neurological disorder (two cases had history of prior stroke). Tissues collected within a post-mortem interval (PMI) less than 48 hours were included. At the time of autopsy, brains were sectioned coronally, and samples of middle frontal gyrus (alternating between left and right sides in the absence of gross abnormalities) were collected and frozen at -80°C. Frozen middle/superior frontal gyrus (Brodmann area 8) controls were obtained from the NIH NeuroBioBank (Harvard Brain Tissue Resource Center/HBTRC and the University of Miami’s Brain Endowment Bank). Controls were selected to be age- and sex-matched to a COVID-19 case and were categorized as unaffected controls (with no known psychiatric or neurological condition) in the NIH NeuroBioBank system. All controls were collected prior to the COVID-19 outbreak in the United States (before 11/2019) and thus are considered uninfected by SARS-CoV-2. RNA integrity (RIN) and PMI were not significantly different between groups.

Frozen tissue was processed using Biosafety Level 2+ (BL2+) procedures approved by the Beth Israel Deaconess Medical Center (BIDMC) Institutional Biosafety Committee (IBC). Brain tissues were homogenized using Trizol (Thermo Fisher) reagent and RNA extracted by phase separation. Total RNA was quantified by nanodrop (DeNovix DS-11) and TapeStation 4200 (RNA Screen Tape; Agilent Technologies, Inc).

### Library construction and RNA-sequencing

450 ng of total RNA was used for library preparation via the KAPA RNA HyperPrep kit with RiboErase (HMR; Roche) according to the manufacturer’s recommendations. Briefly, hybridization with hybridization oligos (HMR) was performed at 95 °C for 2 min followed by rRNA depletion using RNAse H which was performed at 45 °C for 30 min. Following rRNA cleanup via KAPA pure beads, DNase digestion was performed at 37 °C for 30 min followed by cleanup, RNA elution, fragmentation (6 min at 94°C for samples with RIN ≥ 7 or 5 min at 85 °C for samples with RIN ≤ 7), and priming. First and second strand synthesis and A-tailing were performed according to manufacturer’s recommendations. 1.5 μM KAPA Unique Dual-Indexed (UDI; Roche) adapters were ligated to the second strand synthesis product in the presence of a ligation master mix in a reaction that was performed at 20 °C for 15 min. Following cleanup, all libraries underwent 10 cycles of amplification. Successful library production, quality control, and quantification was assessed using Tapestation (High sensitivity D1000 Screen Tape; Agilent Technologies, Inc). All libraries were pooled together (10 nM concentration) and subjected to two lanes of a NovaSeq 6000.

### RT-qPCR (qPCR)

A total of 400 ng RNA from each sample was processed for cDNA via SuperScript IV Reverse Transcriptase kit (ThermoFisher Scientific) according to the manufacturer’s instructions. All qPCR experiments were performed in a 384-well plate using LightCycler 480 SYBR Green via a Roche LightCycler 480 II PCR system.

To assess the expression of SARS-CoV-2, primers against the SARS-CoV-2 N1 and N2 genes were synthesized (IDT) as recommended by the US Center of Disease Control and Prevention (CDC). The 2019-nCoV_N_Positive Control RUO Plasmid (IDT #10006625) was included as positive control. *ITM2B* (Qiagen) was used for normalization. Primers for *S100A9, RHOBTB3, MYL12A, CALM3, INPP4A, GRIA1, and GRIN3A* were designed to span exon-exon junctions using NIH Primer Blast and synthesized by IDT. qPCR data were analyzed via the 2^-ΔΔCt^ method [36].

### RNA-seq analysis

For assessment of SARS-CoV-2 genome alignment: reads were aligned to the SARS-CoV-2 reference genome (NCBI reference sequence NC_045512.2) using bowtie2 v2.2.9 with options “-X 1000 --no-mixed”. Analysis of RNA-seq of Calu-3 infected samples from Blanco-Melo et al. [37] were included as positive controls with default bowtie2 parameters.

For assessment of differential gene expression: raw sequencing reads were aligned to a reference transcriptome generated from the Ensembl v104 human transcriptome with salmon v1.4.0 using options “--seqBias --useVBOpt --gcBias --posBias --numBootstraps 30 -- validateMappings”. Length-scaled transcripts per million were acquired using the tximport v1.18.0 function, and log2 fold changes and false discovery rates (FDR) were determined by DESeq2 v1.30.1 in R. t-stochastic neighboring embedding analysis was performed using Rtsne v0.15, with counts transformed by the varianceStabilizingTransformation (VST) function from DESeq2. Heatmaps were generated using pheatmap v1.0.12 using VST-transformed counts, with further scaling across samples. For full cohort analysis, age/sex-matching was used as a covariate. For cohort subset analyses, no additional covariates were used.

### Gene set enrichment analysis

Signed -log10 FDRs from DESeq2 analyses were used to rank genes for gene set enrichment analysis via fgsea v1.16.0, filtering out genes with an FDR < 0.5. Public gene sets used for analyses: Gene Ontology Biological Processes (GO.db v3.12.1), KEGG (KEGGREST v1.30.1), and ReactomeDB (reactome.db v1.74.0) pathway to gene mappings from fgsea via the “reactomePathways” function. For enrichment analyses, Ensembl gene IDs were matched with corresponding gene symbols and Entrez IDs via biomaRt v2.46.3.

### Brain aging-regulated molecular signatures

We leveraged previously published aging-regulated differentially expressed gene (DEG) set data generated in five independent cohorts [14, 21, 38-41]. Lu et al. (2004) performed a broad-spectrum gene expression analysis (Affymetrix Human Genome U95Av2) of human prefrontal cortex from 30 individuals 26-106 years old and determined age-regulated genes based on a comparison of individuals ≤ 42 vs individuals ≥ 73 years old [14]. Loerch et al. (2008) performed a genome-wide gene expression analysis (Affymetrix Human Genome U133plus 2.0) of human prefrontal cortex from 28 individuals 24-94 years old and determined age-regulated genes based on a comparison of individuals ≤ 40 vs individuals ≥ 70 years old [21]. For three additional human patient cohorts, we used differentially expressed genes as performed by [42] in which gene expression data from individuals 85+ years old were compared to gene expression data of younger individuals, including only individuals with annotated normal cognitive function. Those cohorts include 1) the Gibbs et al. cohort [38] in which gene expression data (Illumina HumanRef-8 Expression BeadChips) from the frontal cortex of 37 individuals were analyzed and DEGs estimated by comparing individuals 85+ to individuals 55-80 years old [42]; 2) the ROSMAP cohort, part of the Religious Order Study (ROS) and Rush Memory and Aging Project (MAP; ROSMAP) at the Rush Alzheimer’s Disease Center [40, 41, 43], in which RNA-seq data from the dorsolateral prefrontal cortex of 117 individuals were analyzed and DEGs estimated by comparing individuals 85+ to individuals 70-80 years old [42]; and 3) the Common Mind Consortium (CMC) cohort [39] in which RNA-seq data from the dorsolateral prefrontal cortex of 155 individuals were analyzed and DEGs estimated by comparing individuals 85+ to individuals 60-80 years old [42].

### Statistical analysis

No statistical methods were used to predetermine sample size. The experimental groups were not randomized. Where possible, samples were processed together using deidentified numbers (RNA-seq library preparation, reverse transcription prior to qPCR, and qPCR). For qPCR and RNA-seq analyses, blinding was not possible as all changes had to be matched with corresponding controls. qPCR data were analyzed with a two-tailed t-test via GraphPad Prism 9. RNA-seq statistical analyses were performed in R v4.0.4 [44].

## Data availability

RNA-seq fastq files generated for this study are available through GEO with accession number GSE188847. Raw Calu-3 .fastq RNA-seq files from Blanco-Melo et al. are available through GEO (https://www.ncbi.nlm.nih.gov/geo/query/acc.cgi?acc=GSE147507) with accession numbers GSM4462348-GSM4462353 [37].

The SARS-CoV-2 genome was obtained from https://www.ncbi.nlm.nih.gov/nuccore/1798174254. The Ensembl v104 human reference transcriptome was obtained from http://ftp.ensembl.org/pub/release-104/fasta/homo_sapiens/cdna/Homo_sapiens.GRCh38.cdna.all.fa.gz. Gene Ontology (http://geneontology.org/) was queried from org.Hs.eg.db v3.12.0 in R. Reactome pathway annotations (https://reactome.org/) were obtained via the “reactomePathways” command in R package “fgsea”: https://bioconductor.org/packages/release/bioc/html/fgsea.html. KEGG hsa pathway annotations (https://www.genome.jp/kegg/) were obtained using the KEGGREST v1.30.1 API in R (https://www.bioconductor.org/packages/release/bioc/html/KEGGREST.html).

---

**Supplementary Figure 1.**
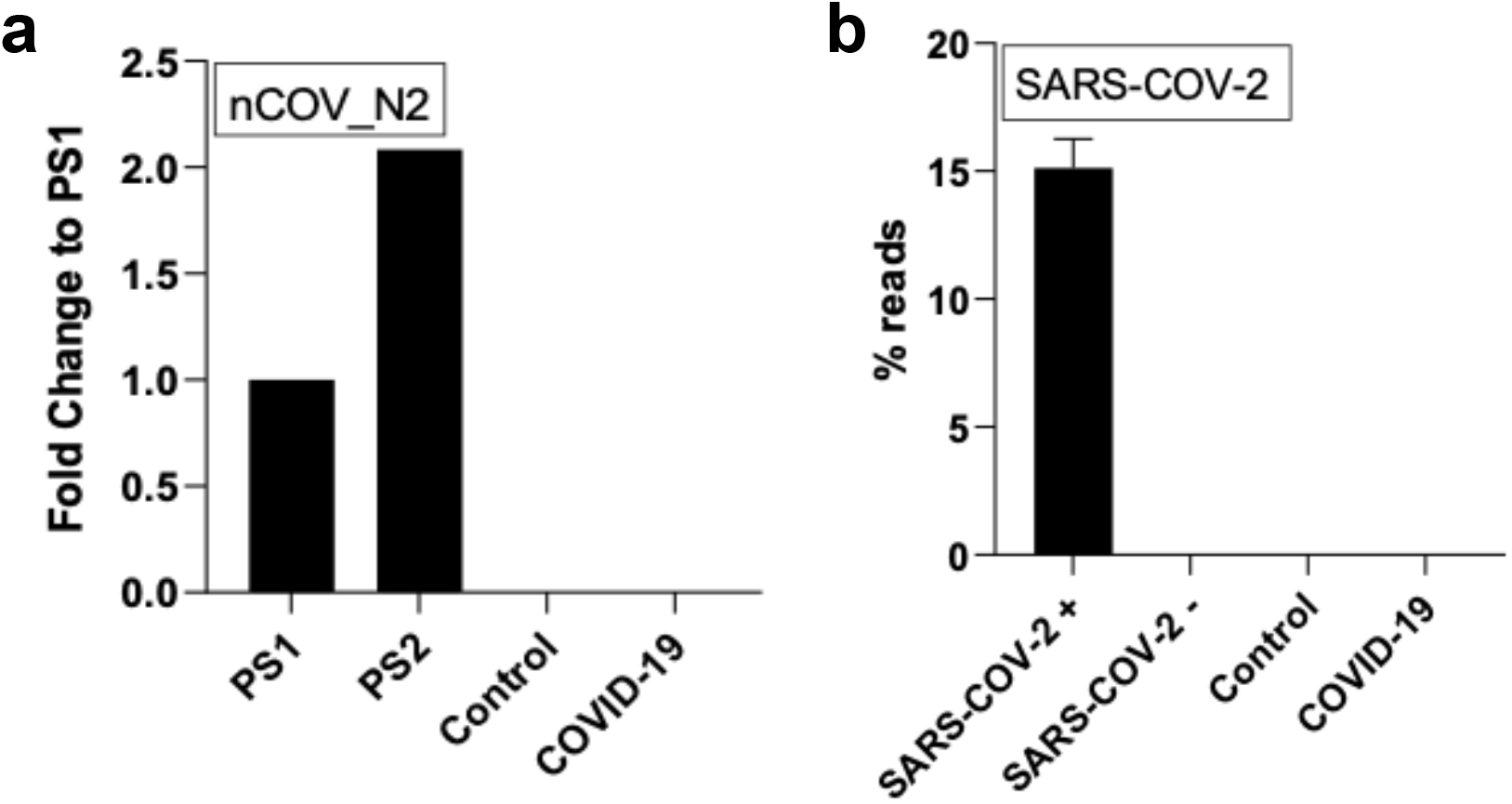
Assessment of SARS-CoV-2 virus in the postmortem human frontal cortex and expression of SARS-CoV-2 brain entry-related genes. **a**. qPCR assessment of SARS-CoV-2 viral RNA in the frontal cortex using the nCOV_N2 primer set. PS1 and PS2 correspond to the 2019-nCoV_N_Positive Control RUO Plasmid (IDT) at concentrations of 1,000 and 2,000 copies/μl, respectively (a technical duplicate/concentration was used to estimate the corresponding mean; for control and COVID-19 samples n=13/group). **b**. Percentages of RNA-seq reads aligning to the SARS-CoV-2 viral genome. SARS-COV-2 + and SARS-COV-2 -correspond to Calu-3 lung cancer cell lines either infected with SARS-CoV-2 virus or mock-infected, respectively [37] (n=3/group for the cell lines and n=12 for the control and COVID-19 groups).

**Supplementary Figure 2.**
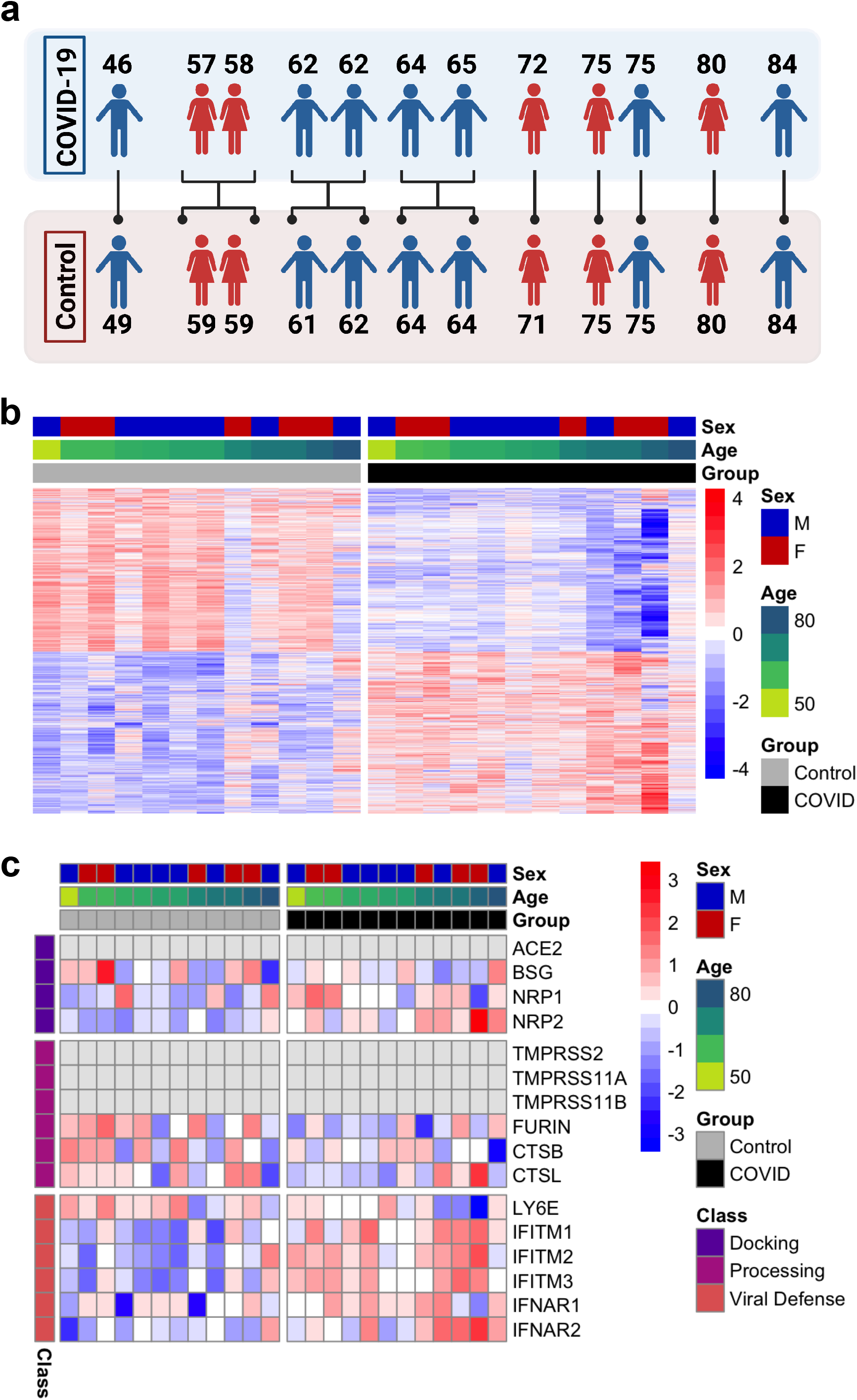
Overview of differential expression patterns in frontal cortex of COVID-19 and uninfected subjects. **a**. Age and sex matching used for differential expression analysis. **b**. Heatmap of relative gene expression levels of all significant DEGs (FDR < 0.05) across COVID-19 and control samples. Color legend, scaled gene expression levels across subjects, normalized via variance-stabilized transformation. **c**. Heatmap of relative expression levels of genes previously implicated in SARS-CoV-2 viral entry in the human brain [8] across COVID-19 and control samples. Color legend, scaled gene expression levels across patients, normalized via variance-stabilized transformation. Light gray cells, no aligned reads or genes filtered out of analysis.

**Supplementary Figure 3.**
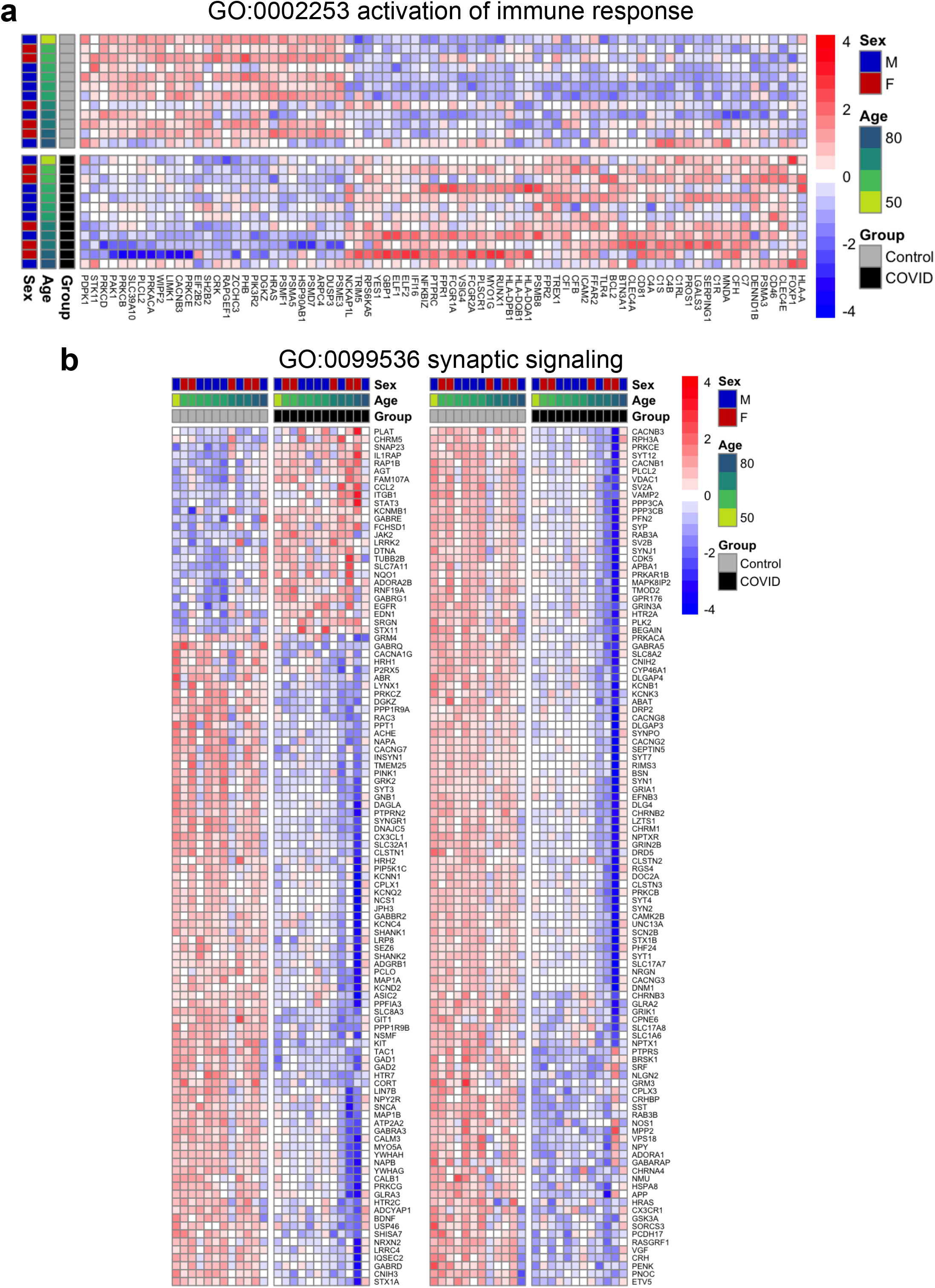
Gene expression patterns of immune response activation and synaptic plasticity pathways. **a** and **b**. Heatmaps of relative gene expression levels of significant DEGs associated with “activation of immune response” (GO: 0002253) (**a**) or “synaptic signaling” (GO: 0099536) (**b**) GO terms across COVID-19 and control samples. Color legend, scaled gene expression levels across subjects, normalized via variance-stabilized transformation.

**Supplementary Figure 4.**
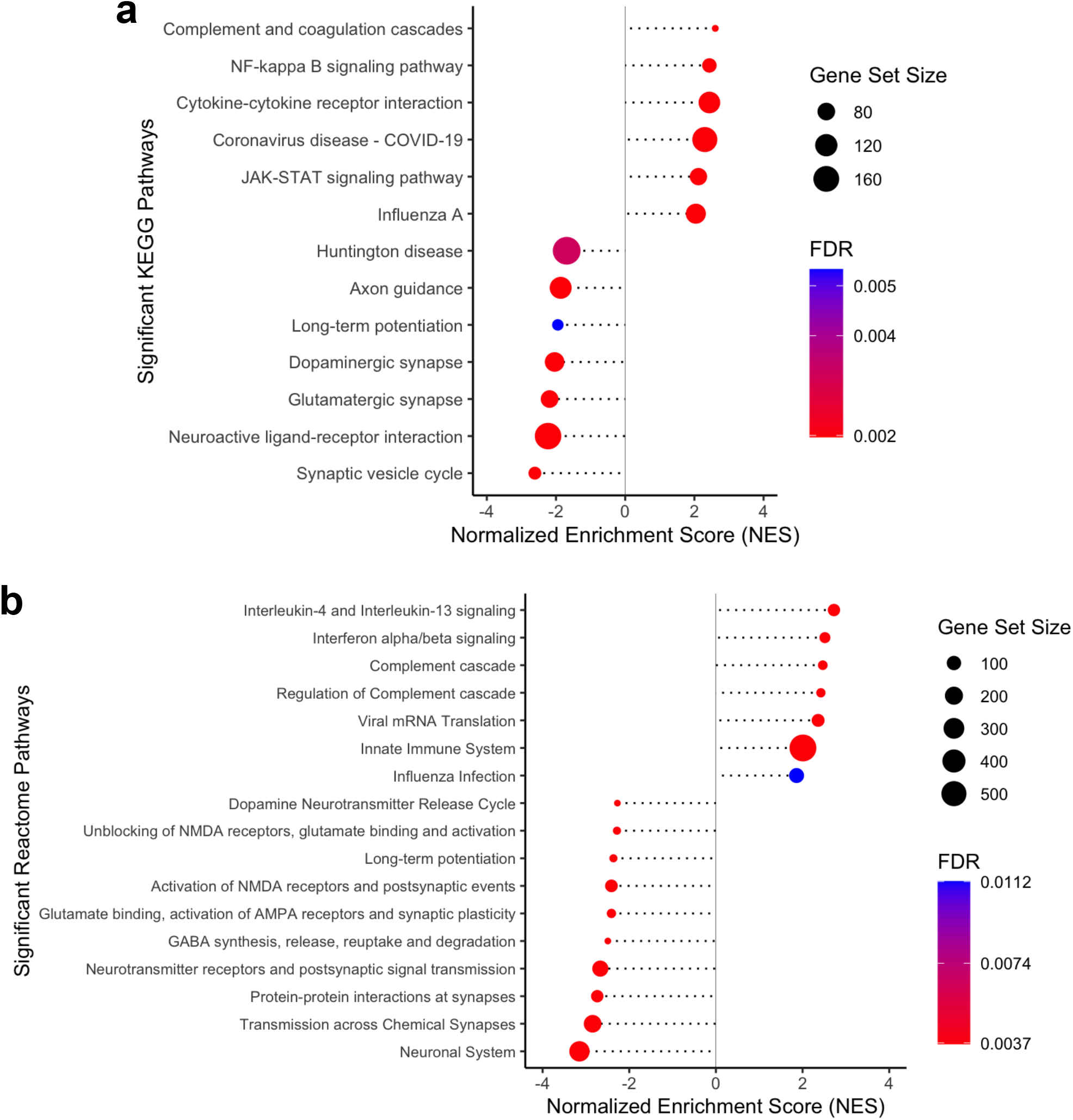
KEGG and Reactome pathway enrichment analyses. **a and b**. Kyoto Encyclopedia of Genes and Genomes (KEGG) (**a**) and Reactome (**b**) pathway enrichment analysis of COVID-19 versus control brain DEGs. Gene ranks were determined by signed -log10 false discovery rates of DEGs. FDR, gene set enrichment analysis false discovery rate.

**Supplementary Figure 5.**
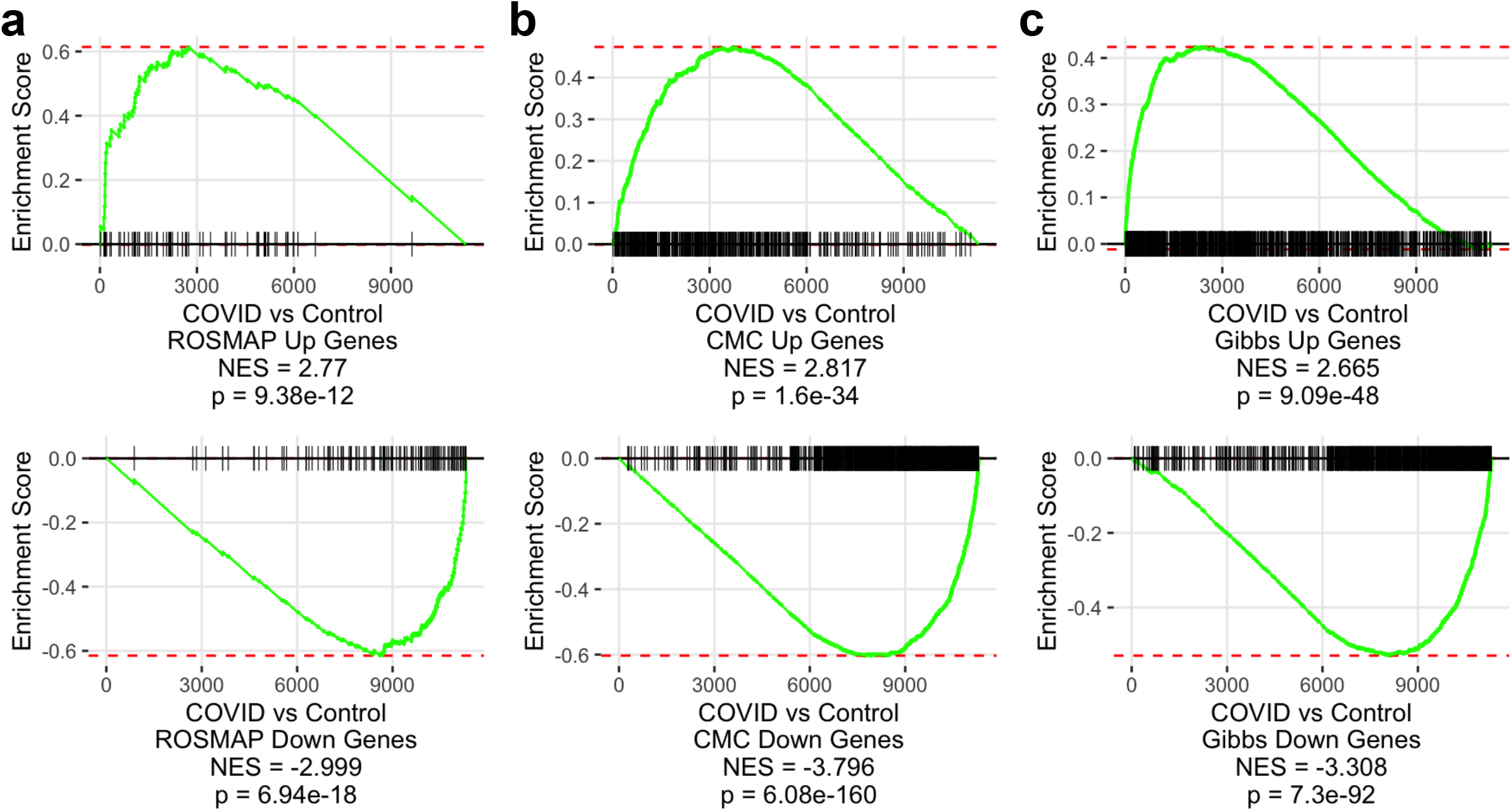
COVID-19 induces molecular signatures found at late age in the human brain. **a-c**. Gene set enrichment analysis of COVID-19 DEGs, using significantly up- (top) or down-regulated genes (bottom) in the ROSMAP (**a**), Common Mind Consortium/CMC (**b**), and Gibbs (**c**) cohorts as gene sets. DEG ranks were assigned by signed -log10 false discovery rates from COVID-19 versus control subject frontal cortex. NES, normalized enrichment score. p, GSEA p-value.

**Supplementary Figure 6.**
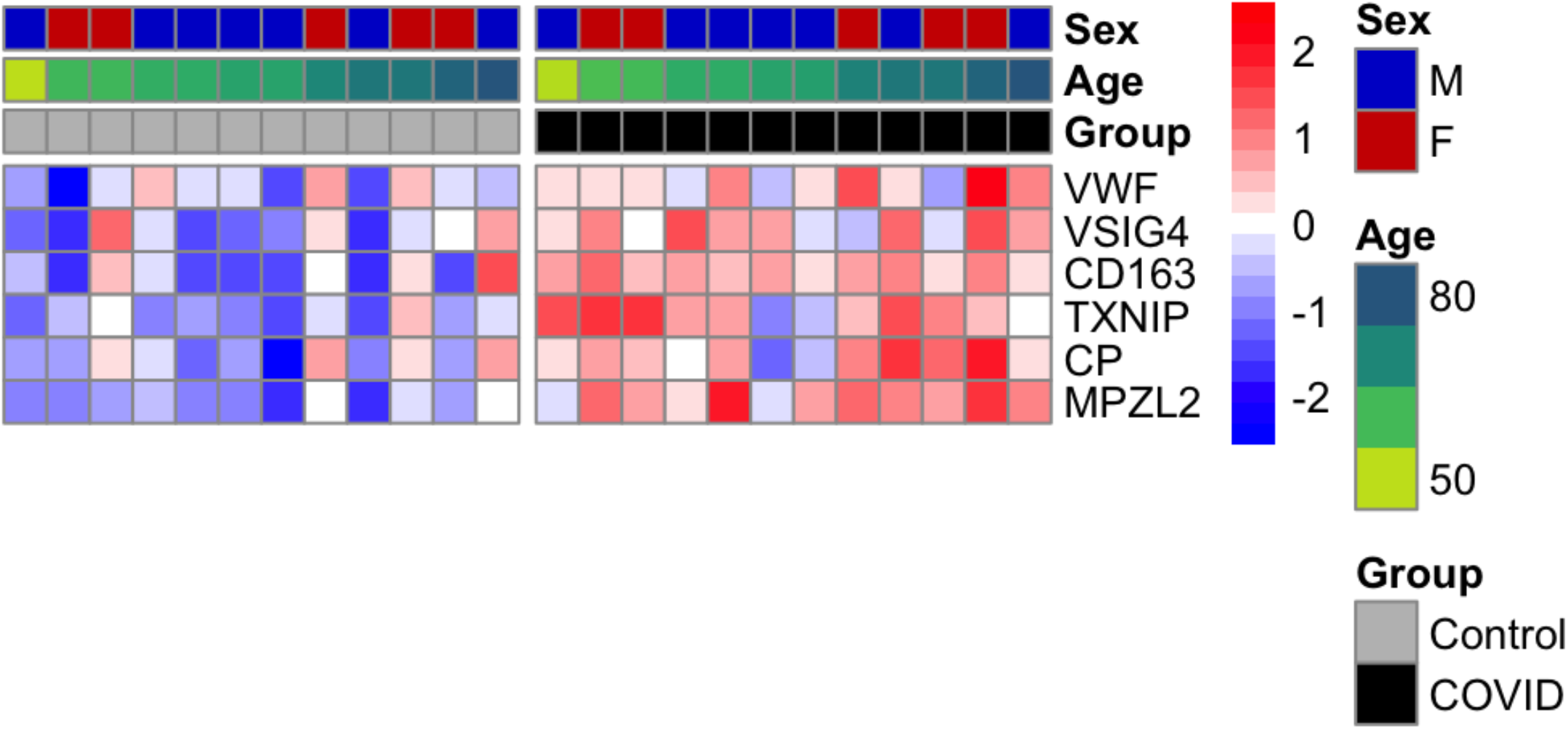
Gene expression patterns of a published brain aging gene signature. Heatmap of relative gene expression levels of a brain aging-distinguishing gene signature from [29] across COVID-19 and control samples. Color legend, scaled gene expression levels across subjects, normalized via variance-stabilized transformation.

**Supplementary Figure 7.**
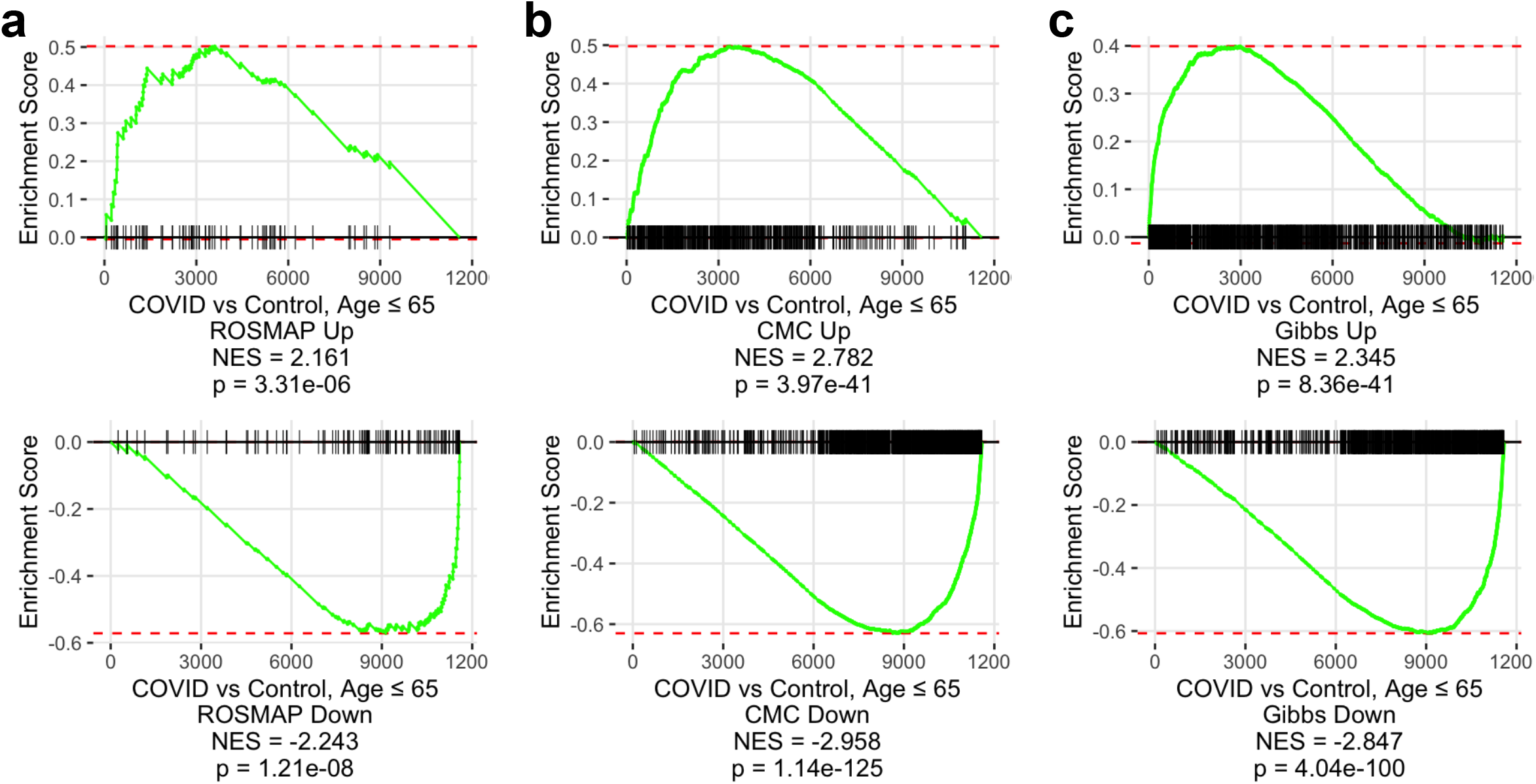
COVID-19 induces molecular signatures found at late age in individuals ≤ 65 years of age. **a-c**. Gene set enrichment analysis of COVID-19 DEGs among individuals ≤ 65 years of age, using significantly up- (top) or down-regulated genes (bottom) in the ROSMAP (**a**), Common Mind Consortium/CMC (**b**), and Gibbs (**c**) cohorts as gene sets. DEG ranks were assigned by signed -log10 false discovery rates from COVID-19 versus control subject frontal cortex among individuals ≤ 65 years of age. NES, normalized enrichment score. p, GSEA p-value.

**Supplementary Figure 8.**
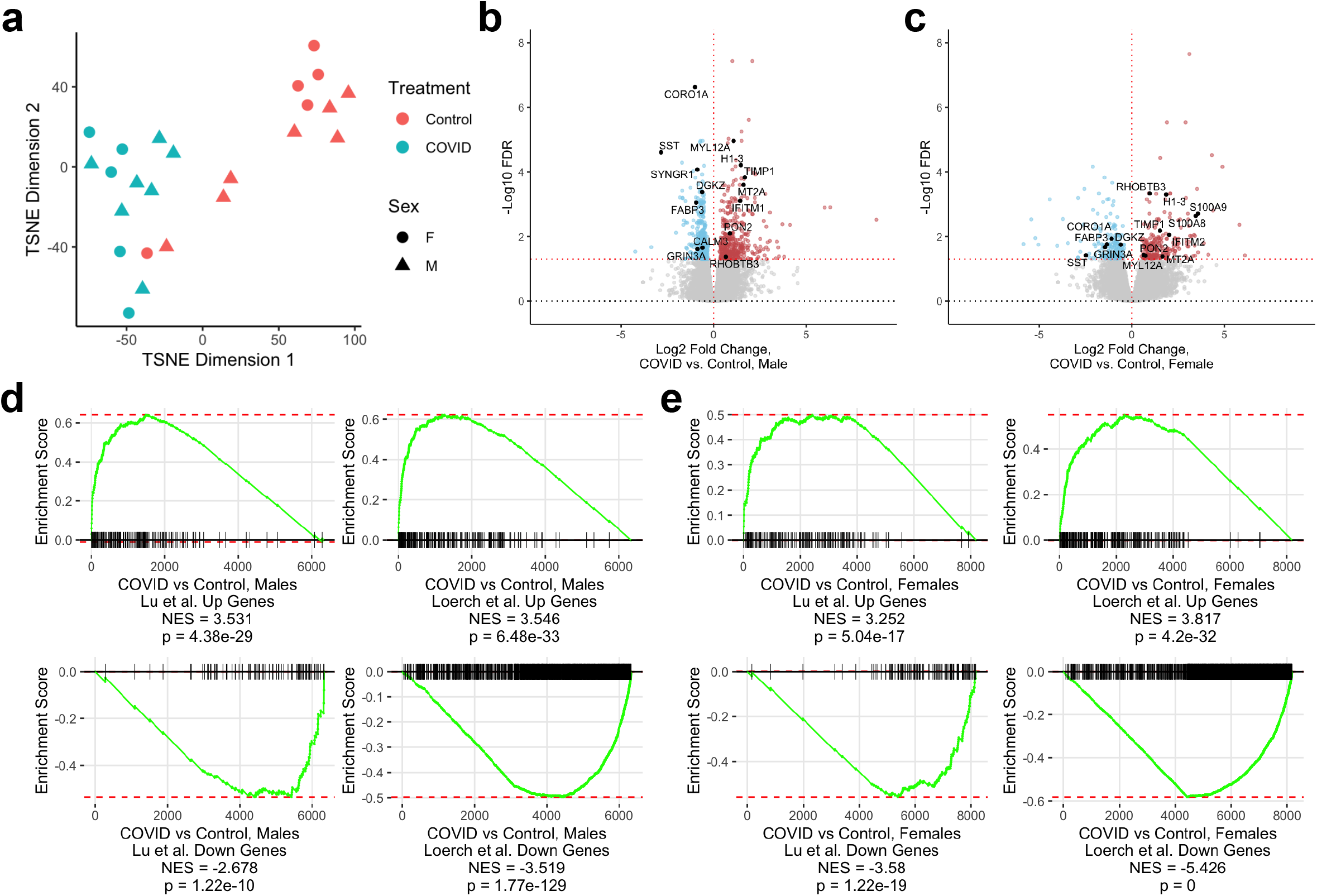
Aging effects of COVID-19 in male and female human brains. **a**. TSNE analysis of frontal cortex transcriptomes from COVID-19 cases and uninfected controls, including sex annotations. **b** and **c**. Volcano plots representing the DEGs of the frontal cortex of COVID-19 cases versus controls among males (**b**, n=7/group) or females (**c**, n=5/group). Red points, significantly upregulated genes among COVID-19 cases (false discovery rate < 0.05). Blue points, significantly downregulated genes among COVID-19 cases. Black points, significant genes with corresponding gene symbols. **d** and **e**. Gene set enrichment analysis of COVID-19 DEGs among males (**d**) and females (**e**), using significantly up- (top) or down-regulated genes (bottom) in the Lu et al. (2004) (left) and Loerch et al. (2008) (right) cohorts as gene sets. DEG ranks were assigned by signed -log10 FDR values from COVID-19 versus control brains among males and females. NES, normalized enrichment score. p, GSEA p-value.

## Notes

### Competing Interest Statement

The authors have declared no competing interest.

### Author Declarations

Post-mortem brain tissue specimens from individuals with severe COVID-19 were collected through an excess tissue waived consent protocol approved by the Mass General Brigham Institutional Review Board.

